# An explainable multimodal framework for exploring depression-associated speech representations in Alzheimer’s disease

**DOI:** 10.64898/2026.09.29.26364278

**Authors:** Hanein Hassan, Dhruv Patel, Hamza Aagela, Anju Johnson

## Abstract

Depression in Alzheimer’s disease (AD) is clinically challenging to characterise because affective and cognitive manifestations frequently overlap with the underlying neurodegenerative condition. This study investigates depression-related speech patterns in AD using an explainable multimodal framework that integrates linguistic and acoustic features from elicited picture-description speech. To address the absence of directly observed depression labels in the target AD cohort, we employ a source-informed cross-dataset transfer strategy. Unsupervised clustering identified two distinct speech sub-phenotypes within the AD cohort. By comparing these unsupervised representations to a depression-labelled source corpus, one cluster demonstrated mathematical proximity (measured via cosine similarity in a shared latent space) to the source depression centroid. We explicitly acknowledge that this proximity reflects geometric similarity in a shared latent space, which may encapsulate overlapping domain characteristics alongside affective variance. To validate the structural integrity and separability of these discovered clusters, a proxy supervised classification task was employed. The retained multimodal artificial neural network (ANN) integrated information from both modalities and achieved 92.73% cluster separability accuracy. This demonstrates that the representation-derived two-cluster partition was highly learnable on the independent internal hold-out test set; it does not establish clinical validity, longitudinal stability, or diagnostic validity of the identified sub-phenotypes. For interpretability, Shapley Additive exPlanations (SHAP) analysis was utilised to extract feature attributions and organise influential acoustic and linguistic features into a multimodal taxonomy. Furthermore, a Multimodal SHAP Attribution Magnitude (MSAM) score was derived as an exploratory model-based attribution score rather than a clinically validated severity measure. Ultimately, this study provides an interpretable computational baseline for characterising depression-related speech variance in AD, establishing a structural foundation for future prospective validation using independently labelled clinical cohorts.

**Author Summary:** Depression can be difficult to recognise in people with Alzheimer’s disease because changes in mood, motivation, language, and behaviour may overlap with symptoms of cognitive impairment. Speech analysis offers a possible way of studying these changes, but datasets containing both Alzheimer’s disease and independently verified depression labels are limited. We therefore explored whether speech representations from an Alzheimer’s disease cohort could be compared with a separate depression-labelled speech dataset. We used acoustic and linguistic features to identify groups of participants with similar speech characteristics and then examined whether these groups could be reliably reproduced by supervised machine-learning models. The best multimodal model reproduced the representation-derived grouping with 92.73% accuracy on an independent participant-level test set. Importantly, this result does not demonstrate that the participants had depression, because depression was not independently assessed in the Alzheimer’s disease cohort. Instead, the study provides an exploratory computational framework for studying cross-corpus speech similarities and identifies features that contributed to the learned speech representations. Prospective studies with clinically assessed depression status are needed before such methods can be considered for clinical use. Such studies should also examine whether the observed speech patterns remain informative after accounting for cognitive impairment, age, medication, speaking task, and recording conditions. This work therefore represents a hypothesis-generating computational step rather than a diagnostic tool.

## Introduction

Depression and dementia are prevalent disorders among older adults that frequently coexist, creating substantial challenges for patient management and quality of life [1]. While depression is potentially treatable through pharmacological and psychological interventions, dementia, particularly Alzheimer’s disease (AD) [2], is characterised by progressive neurodegenerative decline. Differentiating depression-related symptoms from manifestations of AD is clinically important, yet inherently difficult due to overlapping manifestations such as apathy, sleep disturbances, changes in weight, reduced motivation, and impaired concentration [3]. Such symptom overlap can complicate the recognition of depression when it occurs in the context of established cognitive impairment.

Beyond differential diagnosis, recognising depression-related manifestations in individuals with AD is clinically important because comorbidity has been associated with adverse patient and caregiver outcomes [4]. The coexistence of the two conditions has been associated with accelerated cognitive decline, earlier institutionalisation, increased caregiver burden, and higher mortality [5, 6]. Despite these consequences, depression-related symptoms in people with AD may remain difficult to recognise, particularly when assessment relies heavily on subjective clinical judgement.

Recent advances in speech processing and artificial intelligence have demonstrated that depression and dementia are associated with measurable changes in both acoustic and linguistic behaviour [7]. However, comparatively limited research has investigated depression-related speech characteristics specifically within an AD population using a joint multimodal analysis. This gap is important because unimodal representations may capture only part of the behavioural signal associated with depression in the presence of neurodegenerative impairment.

In this study, we develop an explainable multimodal speech framework to explore depression-related variance in AD. Acknowledging the lack of ground-truth comorbidity labels in our target cohort, we do not attempt direct diagnostic classification. Instead, we propose a pipeline centred on unsupervised phenotype discovery, pseudo-label alignment, and structural stability evaluation. The resulting pseudo-labels are used to evaluate text-only, audio-only, and multimodal representations with classical machine-learning and neural models. Cross-validation and independent hold-out evaluation are employed to assess robustness, while SHAP-based analysis is used to identify influential acoustic and linguistic features.

**The main contributions of this study are as follows:**

- Develop an exploratory multimodal speech framework that integrates acoustic and linguistic representations to map depression-related patterns within Alzheimer’s disease.
- Establish an exploratory cross-dataset alignment strategy that maps geometric similarities between an unlabelled AD cohort and a distinct depression-labelled source, serving as a heuristic proxy rather than a definitive clinical classifier.
- Utilise supervised machine learning explicitly to evaluate the non-linear separability and mathematical stability of the discovered sub-phenotypes, rather than making unsupported diagnostic claims.
- Deploy SHAP-based explainability across the experimental settings to quantify geometric feature contributions, thereby organising influential acoustic and linguistic characteristics into an interpretable multimodal feature-attribution taxonomy.
- Reframe the supervised classification task as a direct measure of the learnability of the unsupervised Gaussian Mixture Model (GMM) latent partition, ensuring the evaluation avoids methodological circularity and focuses on algorithmic stability rather than clinical construct validity.

The remainder of this paper is organised as follows. The Related work section reviews speech-based depression and dementia analysis. The Materials and methods section describes the proposed cross-dataset transfer, pseudo-labelling, multimodal representation, classification, and explainability framework. The Results section presents the experimental evaluations, the Discussion interprets the findings in relation to previous work and the methodological limitations of the study, and the Conclusion summarises the principal implications and directions for future research.

## Related work

Speech contains acoustic and linguistic characteristics that have been investigated as potential markers of depression and cognitive impairment. However, these abnormalities may not be readily perceptible to human observation and require technical integration for their detection [8]. Furthermore, research has demonstrated that both acoustic and linguistic features have been investigated as complementary representations of speech changes associated with neurological and affective conditions. Ehghaghi et al. [9] evaluated the effectiveness of clustering methods for distinguishing between depression and dementia using human speech data. An innovative aggregated dataset was introduced, combining narrative speech data from various conditions, including AD, mild cognitive impairment, healthy controls, and depression. This study compared linear and nonlinear clustering approaches, demonstrating that nonlinear clustering techniques more effectively differentiate between distinct disease clusters. The authors [3] employed linguistic features to examine the influence of depression on the detection of AD by categorising participants into depressive and nondepressive groups. In this experiment, the optimal model achieved a classification accuracy of 65.8% using the Logistic Regression classifier. This underscores the complexity and context-dependence of both detection tasks (AD vs. non-AD and depressive vs. non-depressive).

The objective of this study [10] was to develop a foundational methodology for utilising Artificial Neural Networks (ANN) to categorize patients with AD into depressive and non-depressive groups through speech analysis. The classification model reported classification accuracies of 72.5%–77.1%. These findings indicate that ANN holds potential to assist the medical community in future research initiatives.

Nevertheless, these studies continue to encounter difficulties in delivering comprehensive, clinically actionable insights that effectively bridge the gap between machine learning predictions and medical decision-making.

## Materials and methods

The proposed framework consists of five essential stages: dataset preprocessing, feature extraction and selection, multimodal feature fusion, structural cluster validation (via machine learning and deep learning), and SHAP-based feature attribution. The specifics of each stage are detailed as follows.

### Dataset

In this study, we used two complementary speech data corpora, DementiaBank and DAIC-WOZ, to investigate speech representations associated with depression in a source corpus and their geometric correspondence to an Alzheimer’s disease cohort while addressing the cognitive deficits associated with AD.

### DementiaBank

This clinical database is widely used in research and includes audio recordings and transcriptions of individuals diagnosed with cognitive disorders, along with age-matched healthy control speakers [11]. These recordings were sourced from participants who engaged in the ’Cookie Theft’ picture description task [12]. The original dataset comprises 302 participants. Following stringent quality control, 29 recordings were excluded due to severe acoustic distortion or incomplete transcriptions, resulting in a final retained cohort of 273 participants. The demographic and clinical diagnostic breakdown of this cohort (including AD, Mild Cognitive Impairment, and healthy controls) is detailed in retained cohort of 273 participants. The demographic and clinical diagnostic breakdown of this cohort (comprising AD and Mild Cognitive Impairment) is detailed in Table 1. Before analysis, all audio recordings and transcriptions were processed using the following predefined steps to remove interviewer speech and to consolidate task-level recordings into a single concatenated file for each participant.

**Table 1.** Participant characteristics and diagnostic composition for the DementiaBank target cohort.

| Variable | AD | MCI | Total |
| --- | --- | --- | --- |
| $N$ (Participants) | 240 | 33 | 273 |
| Age (Avg.Range) | 49-90 |  | 49-90 |
| Sex (Female, $n$) | — | | 169 |
| Sex (Male, $n$) | — | | 104 |
| *Note: Age and sex demographics reflect the combined retained cohort.* |  |  |  |

### DAIC-WOZ

This dataset is specifically designed for the assessment of depression through the analysis of spoken language [13]. DAIC-WOZ contains interview-based speech recordings accompanied by depression-related clinical assessments. The present analysis used the training and development partitions comprising 142 participants, for whom PHQ-8 scores were available. Notably, this source cohort was not recruited as an Alzheimer’s disease cohort. It was therefore used as a depression-labelled reference corpus rather than as a matched control population, and differences between the source and target cohorts were treated as potential sources of domain shift. The experiments utilised the training and development sets, comprising 142 subjects (63 males and 79 females) with an average PHQ-8 score of 6.67 [14].

To establish an exploratory, data-driven anchor for our target AD cohort without falsely claiming a clinical ’ground truth’, we employed a cross-dataset feature alignment strategy. Specifically, the acoustic and linguistic representation of the depression-labelled source subset was used as a heuristic reference for comparison with the DementiaBank target clusters. In the corresponding target-domain representation, the heuristic alignment identified one of the two target clusters as more similar to the source depression representation. The size of the selected cluster was representation-dependent and is therefore reported separately for the text, acoustic, and fused analyses rather than treated as a common clinical pseudo-label. We note that given the fundamental domain differences between the corpora, namely task design (picture description versus clinical interview), demographic profiles, and recording conditions, this alignment captures mathematical proximity to a depression-associated representation rather than an isolated clinical ground truth unaffected by domain variables.

### Feature extraction and selection

Acoustic and linguistic features were extracted from the participant-level speech recordings and corresponding transcripts. The acoustic representation captured energy, spectral, voice-quality, and cepstral characteristics, whereas the linguistic representation combined TF-IDF features with manually derived lexical and sentiment-related measures. Table 2 summarises the feature groups used throughout the experiments.

**Table 2.** Summary of extracted features.

| Feature Category | Feature Type | Description |
| --- | --- | --- |
| Acoustic | RMS | Root-mean-square signal energy |
|  | ZCR | Zero-crossing rate |
|  | Centroid | Spectral centre of mass [15] |
|  | Roll-off | Frequency below which a specified proportion of spectral energy is concentrated [16] |
|  | Jitter | Relative short-term variation in estimated fundamental-period duration |
|  | Shimmer | Relative short-term variation in frame-level amplitude |
|  | MFCC<sub>0</sub>–MFCC<sub>12</sub> | Thirteen Mel-frequency cepstral coefficients representing spectral characteristics [16] |
| Linguistic | TF-IDF | Term-level lexical representation |
|  | Word Count | Total number of tokens [17] |
|  | Sentence Count | Number of detected sentences [18] |
|  | Average Sentence Length | Mean number of words per sentence [19] |
|  | TTR | Type-token ratio representing lexical diversity [20] |
|  | Polarity | Sentiment polarity representing emotional valence [18] |
|  | Subjectivity | Subjective versus objective language tendency [21] |

The acoustic representation consisted of 19 features, extracted after downsampling all audio to a uniform 16 kHz sample rate to ensure cross-corpus consistency. Frame-level extraction utilised a 25 ms frame length with a 10 ms hop length, applying a standard Hamming window. To reduce the contribution of low-energy and non-speech frames, frames below a predefined RMS-energy threshold were excluded prior to participant-level aggregation. The remaining frame-level descriptors RMS, ZCR, spectral centroid, spectral roll-off, local absolute jitter, local relative shimmer, and 13 Mel-frequency cepstral coefficients (extracted using 40 Mel bands between 20 Hz and 8000 Hz) were aggregated to the participant level by calculating the statistical mean across all active voiced frames. Missing values resulting from severely distorted audio segments were imputed using the respective feature mean from the training partition. The linguistic pipeline began with standardising the transcripts through lowercasing, punctuation removal, and tokenisation using a standard word tokeniser. Domain-agnostic stop-words were removed, and the remaining vocabulary was reduced to base forms using WordNet lemmatisation. The TF-IDF weighting was configured to capture both unigrams and bigrams, applying L2 normalisation and a minimum document frequency of five to eliminate highly sparse idiosyncratic terms. The TF-IDF vocabulary was limited to a maximum of 500 features using the vectoriser’s feature-selection procedure after application of the specified document-frequency constraints. Sentiment polarity and subjectivity were calculated using a pre-trained lexicon-based sentiment model.

Combined with the lexical descriptors (e.g., word count, TTR), this yielded 506 textual features. These representations were retained separately before modality-specific standardisation.

All preprocessing operations that estimate parameters from the data, including feature scaling and dimensionality reduction, were strictly restricted to the corresponding training partition during model validation to prevent data leakage. The fitted transformations were subsequently applied to the held-out fold or independent test set without refitting.

### Multimodal feature representation and fusion

Rather than directly concatenating the full acoustic and linguistic feature spaces, the two modalities were processed independently to reduce dimensional imbalance and preserve modality-specific characteristics. Let **x***_a_ ∈* R*^da^* and **x***_t_ ∈* R*^dt^* denote the acoustic and linguistic feature vectors, respectively. After modality-specific standardisation (scaling to zero mean and unit variance based exclusively on the training fold), each representation was projected into a lower-dimensional latent space using principal component analysis (PCA). To reduce dimensionality while retaining 95% of the variance observed in the training representation, the number of retained components was set to capture 95% of the cumulative explained variance. Crucially, the PCA transformation fitted on the target training data was applied identically to the source representation, allowing the source representation to be transformed into the corresponding target-derived coordinate system before cross-corpus comparison:

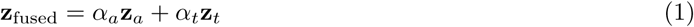

The modality-specific latent representations were subsequently combined using weighted feature concatenation. This preserves the separate coordinate systems of the acoustic and linguistic latent spaces while allowing each modality to contribute to the fused representation.

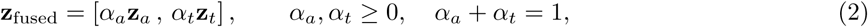

Here, *α_a_* and *α_t_* denote the relative contributions assigned to the acoustic and linguistic representations, respectively. Crucially, these modality weights specify structural contributions to the fused feature space and must not be interpreted as measures of clinical reliability or diagnostic confidence. The resulting multimodal representation was subsequently used for unsupervised clustering and supervised validation. It is imperative to note that to rigorously explore the distinct latent structures of each modality, clustering and pseudo-label generation were performed independently within the text-only, audio-only, and fused latent spaces.

Consequently, the resulting pseudo-label distributions naturally vary across the text, audio, and fusion experiments. Because each modality possesses a uniquely shaped geometric topology, the pseudo-labels serve as modality-specific structural targets rather than a universally fixed clinical label. We strictly caveat that this prohibits a direct one-to-one performance comparison across modalities, as the target class distributions inherently shift across the independent evaluations.

### Unsupervised clustering and pseudo-label generation

Because independent clinical depression annotations were absent for the target AD cohort, unsupervised clustering was strictly utilised as an exploratory mechanism to identify geometric structure within the target-domain representations. A Gaussian Mixture Model (GMM) was fitted to the training representation. To ensure reproducibility, the GMM was configured with full covariance matrices, a convergence tolerance of 1 *×* 10*^−^*^3^, a maximum of 100 iterations, and 10 repeated random initialisations. Candidate values of *k* were evaluated using the Silhouette score. Cluster stability was further evaluated by verifying membership consistency across repeated random seeds. The value of *k* yielding the highest Silhouette score was selected for pseudo-label generation.

These resulting clusters represent abstract speech-pattern groups, not clinically confirmed diagnostic categories. To associate these target-domain clusters with depression-related characteristics, each target cluster centroid was compared with the centroid of the depression-labelled source representation using cosine similarity. The cluster exhibiting the highest similarity was heuristically designated as the ’depression-like’ cluster.

To evaluate the mathematical stability of these discovered groups, the resulting assignments were treated as discrete classes for a downstream proxy classification task. We explicitly state that our supervised classifiers were trained solely to predict these unsupervised cluster assignments. By establishing how accurately a multimodal model separates these clusters, we measure the learnability of the GMM latent partition. This approach quantifies the geometric distinctiveness of the target clusters while actively avoiding the methodological circularity of validating the construct of depression through self-derived pseudo-labels.

### Depression-specific feature transfer and pseudo-label alignment

Let the depression-labelled source corpus be 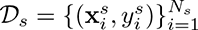, where 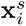 denotes the extracted representation and 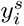 denotes the corresponding source-domain depression label. The target AD cohort is represented as 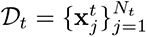, for which independent depression annotations are not assumed.

Rather than learning an explicit parametric domain-alignment function, the proposed transfer procedure constructs a heuristic source depression representation. For the text-based cross-corpus comparison, the source and target corpora were represented in a shared TF-IDF feature space constructed from their intersecting vocabulary. The vocabulary was restricted to the 500 highest-frequency terms satisfying the predefined corpus-level frequency criteria. This procedure ensured dimensional correspondence between the source and target text representations; it did not, however, remove broader differences in task, recording conditions, demographics, or language use between the corpora. The target-domain clusters are then compared with this source representation. Furthermore, for the fusion formulation (Eq 2), an unweighted equal-contribution scheme was adopted (*α_a_* = 0.5, α_t_ = 0.5). Because the preceding PCA transformations effectively normalised the variance scales of both latent spaces, this equal weighting ensures the model is not inadvertently biased toward the modality with a higher intrinsic variance. Let *D_s,_*_+_ denote the depression-labelled subset of the source corpus, defined using the source corpus’s prespecified binary depression label. The centroid of this depression-labelled source representation is then defined as

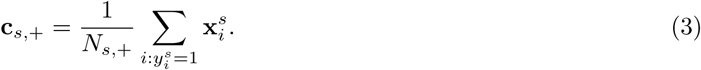

For a target cluster *C_k_*, its centroid is defined as

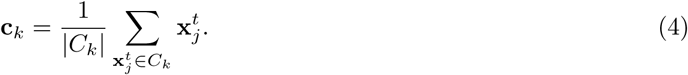

The correspondence between the source depression representation and each target cluster is quantified using cosine similarity:

To rigorously assess alignment, each target cluster was compared against both the source depression centroid (**c***_s,_*_dep_) and the source non-depression centroid (**c***_s,_*_nondep_). The relative mathematical proximity (Δ*_k_*) was quantified as the difference in cosine similarities:

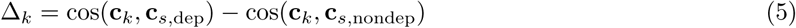

The target cluster yielding the largest positive Δ*_k_* difference was heuristically designated as the depression-like cluster:

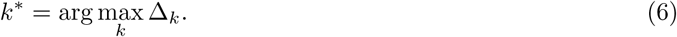

To rigorously assess alignment, each target cluster was compared against both the source depression centroid (**c***_s,_*_dep_) and the source non-depression centroid (**c***_s,_*_nondep_). The relative mathematical proximity (Δ*_k_*) was quantified as the difference in cosine similarities:

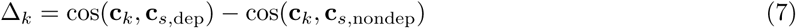

The target cluster yielding the largest positive Δ*_k_* difference was heuristically designated as the depression-like cluster:

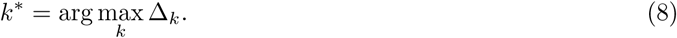

Specifically, the cross-corpus comparison yielded distinct alignment profiles for the target partition. Cluster 1 demonstrated a higher semantic (cosine) similarity of 0.1781 and a lower sentiment distance of 0.4573 relative to the source representations. In contrast, Cluster 0 exhibited a lower semantic similarity of 0.0898 alongside a substantially higher sentiment distance of 0.9665. Because Cluster 1 yielded the stronger mathematical proximity to the depression-associated source representation, it was heuristically designated as the target depression-like cluster.

This selected cluster assignment acts merely as a data-driven anchor. Consequently, all subsequent performance metrics reflect structural feature distinctiveness and cluster separability, not diagnostic efficacy.

### Explainability and exploratory attribution summary

To map the decision boundaries separating our discovered sub-phenotypes, SHAP (using the KernelExplainer) was employed [**?**]. Attributions were calculated on the log-odds output scale using the corresponding training fold as the background reference distribution. Highly correlated MFCC variables were retained to observe holistic spectral attribution, though we acknowledge this distributes importance across correlated dimensions. SHAP values quantify the geometric contribution of individual input features to the model’s structural separation, relative to a reference prediction. The resulting attributions were organised into a multimodal feature-attribution taxonomy to describe model-derived feature behaviour.

As an exploratory attribution summary, we additionally define a Multimodal SHAP Attribution Magnitude (MSAM). The index is introduced here as a study-specific descriptive quantity and is not presented as an established clinical or statistical measure. For an individual instance *x*, the score aggregates the magnitudes of the most influential acoustic and linguistic SHAP values:

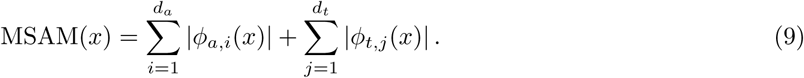

Here, *ϕ_a,i_*(*x*) and *ϕ_t,j_*(*x*) denote the SHAP contributions of the acoustic and linguistic features, respectively, with *d_a_* and *d_t_* denoting the corresponding numbers of features included in the attribution summary. The resulting MSAM provides a unified model-attribution score that summarises the magnitude of influential multimodal features for an individual instance. It is strictly an exploratory interpretability measure and must not be interpreted as a clinically validated severity scale.

### Classification architecture (latent partition learnability)

To compare conventional and neural boundaries, four classical machine-learning models were evaluated: Logistic Regression, Random Forest, Support Vector Machine (SVM), and XGBoost. These models are deployed not as diagnostic classifiers, but as structural validation vehicles to test whether the unsupervised GMM boundaries can be successfully approximated and learned in a non-linear latent space.

For the SVM baseline, a radial basis function (RBF) kernel with *C* = 1.0 was employed. Logistic

Regression and Random Forest were configured with class-balancing strategies, while XGBoost was used as a gradient-boosted tree baseline under the corresponding binary-classification configuration.

For the neural experiments, the multimodal representation was processed using a fully connected artificial neural network (ANN). To ensure reproducibility across all runs, the global random seed was fixed and network weights were initialised using He normal initialisation. The architecture comprised a single hidden layer of either 128 (ANN-128) or 256 (ANN-256) dense units, immediately followed by Batch Normalisation and Dropout regularisation (*p* = 0.3) to heavily constrain model flexibility given the small participant sample. The final layer utilised a sigmoid activation for binary classification. Models were trained using the Adam optimiser with a learning rate of 0.001 and a batch size of 32. To further prevent over-parameterisation from leading to memorisation, training was capped at 100 epochs, with an early stopping patience of 15 epochs monitoring the validation loss. The hyperparameter configuration was locked following the cross-validation phase, ensuring no data leakage into the hold-out evaluations.

### Evaluation metrics (cluster separability)

To quantify the ability of the models to reproduce the unsupervised cluster assignments, we evaluated accuracy, balanced accuracy, macro-averaged F1-score, sensitivity, specificity, ROC-AUC, and PR-AUC. Because the pseudo-label distributions were not necessarily balanced, balanced accuracy and macro-F1 were used alongside accuracy to reduce dependence on the majority class. All metrics were computed on participant-level predictions, and the averaging convention was kept consistent across models and experiments.

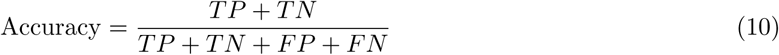

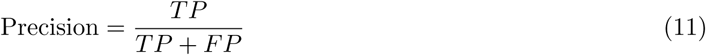

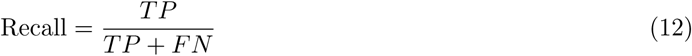

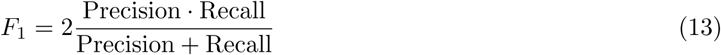

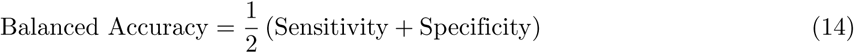

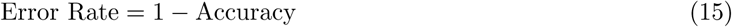

ROC-AUC and PR-AUC were additionally utilised to evaluate the model’s ability to rank the pseudo-labelled instances across decision thresholds. For five-fold cross-validation, each metric was summarised using the mean and standard deviation across folds.

### Validation protocol

Model evaluation followed a participant-level five-fold cross-validation protocol together with an independent hold-out test set [22]. The participant-level design ensured that observations from the same participant were not distributed across training and evaluation partitions.

All data-dependent transformations were fitted exclusively within the corresponding training partition.

These operations included TF-IDF vocabulary and weighting estimation, feature standardisation, PCA, GMM parameter estimation, source-to-target cluster alignment, and classifier fitting. The resulting fitted transformations and models were then applied without refitting to the corresponding validation fold. The final 20% test set was kept untouched until all methodological and model-selection decisions had been completed.

The five-fold procedure was used to estimate model stability across different training and evaluation partitions. Performance metrics were reported as mean *±* standard deviation across the five folds. An independent hold-out test set was retained for the final evaluation after the methodological and modelling choices had been established using the training data and cross-validation procedure.

## Results

Three experimental settings were evaluated to measure the geometric separability of the discovered sub-phenotypes: text-only, audio-only, and multimodal fusion. A participant-level 80/20 split allocated 218 participants for training and 55 for independent structural validation testing. Five-fold cross-validation was used for model selection, with results reported as mean *±* standard deviation. Audio training data were augmented, while the test set remained unchanged. Logistic Regression, Random Forest, SVM, and XGBoost were evaluated, with ANN-128 and ANN-256 additionally assessed for multimodal fusion. Within each training partition, source-informed clustering and pseudo-label assignment were performed using only the training participants. The fitted transformations and GMM parameters were subsequently applied to the corresponding held-out participants without refitting. The source-informed alignment was used only as an exploratory heuristic. Experiments were executed using Google Colab, Python 3.10, TensorFlow, Scikit-learn, SHAP, and an NVIDIA T4 GPU.

### Clustering analysis (structural discovery)

Before proxy classification, an unsupervised clustering analysis was conducted to investigate the underlying geometric structure and non-linear separability of the acoustic-linguistic latent space within the Alzheimer’s data. The optimal number of clusters was determined using the Silhouette Score.The evaluated cluster configurations yielded the following scores: *k* = 2 (0.487), *k* = 3 (0.482), *k* = 4 (0.429), and *k* = 5 (0.305). The highest observed score was obtained for *k* = 2, indicating that the two-cluster solution provided the strongest separation among the candidate solutions evaluated in this analysis. This result provided exploratory support for a two-cluster representation within the analysed AD cohort and motivated the subsequent heuristic comparison of the two target clusters with the depression-labelled source representation.

### Text-based experiment

The text-only experiment used 506 textual features, comprising 500 TF-IDF dimensions and six participant-level linguistic descriptors: word count, sentence count, average sentence length, type token ratio (TTR), sentiment polarity, and subjectivity. A participant-level split allocated 218 participants to training and 55 to the independent test set. The target labels were generated through the pseudo-labelling procedure described in the Materials and methods section and explicitly represent model-derived structural groups rather than independently verified clinical diagnoses.

Key findings:

- Strongest structural boundary: As detailed in Table 3, Random Forest achieved the highest hold-out separability accuracy (87.27%) and F1-score (0.8770). The confusion matrix (Fig 1) and ROC curve (Fig 3) further illustrate this model’s discriminative boundary on the test set. Additionally, it produced the strongest cross-validated accuracy (81.34 ± 7.83%) and F1-score (81.49 ± 7.08%), as compared in Fig 2.
- Strong ranking performance of SVM: Although SVM produced a lower threshold-based accuracy (76.36%), it achieved the highest hold-out ROC-AUC (0.9592) and cross-validated ROC-AUC (0.9761 ± 0.0196) (Table 3), indicating strong discriminative ranking of the pseudo-labels.
- Class imbalance relevance: The difference between Accuracy, F1, and Balanced Accuracy indicates that the structural boundary mapping is affected by the unequal distribution of the pseudo-labelled groups.
- Interpretability: The distribution of term-weight values across textual instances is visualised in the TF-IDF heatmap (Fig 4). Correspondingly, SHAP analysis (Fig 5) identified sentence count, TTR, subjectivity, and selected pronoun-related features among the influential textual variables, highlighting how discourse structure and lexical properties mathematically divide the sub-phenotypes.

**Fig 1.**
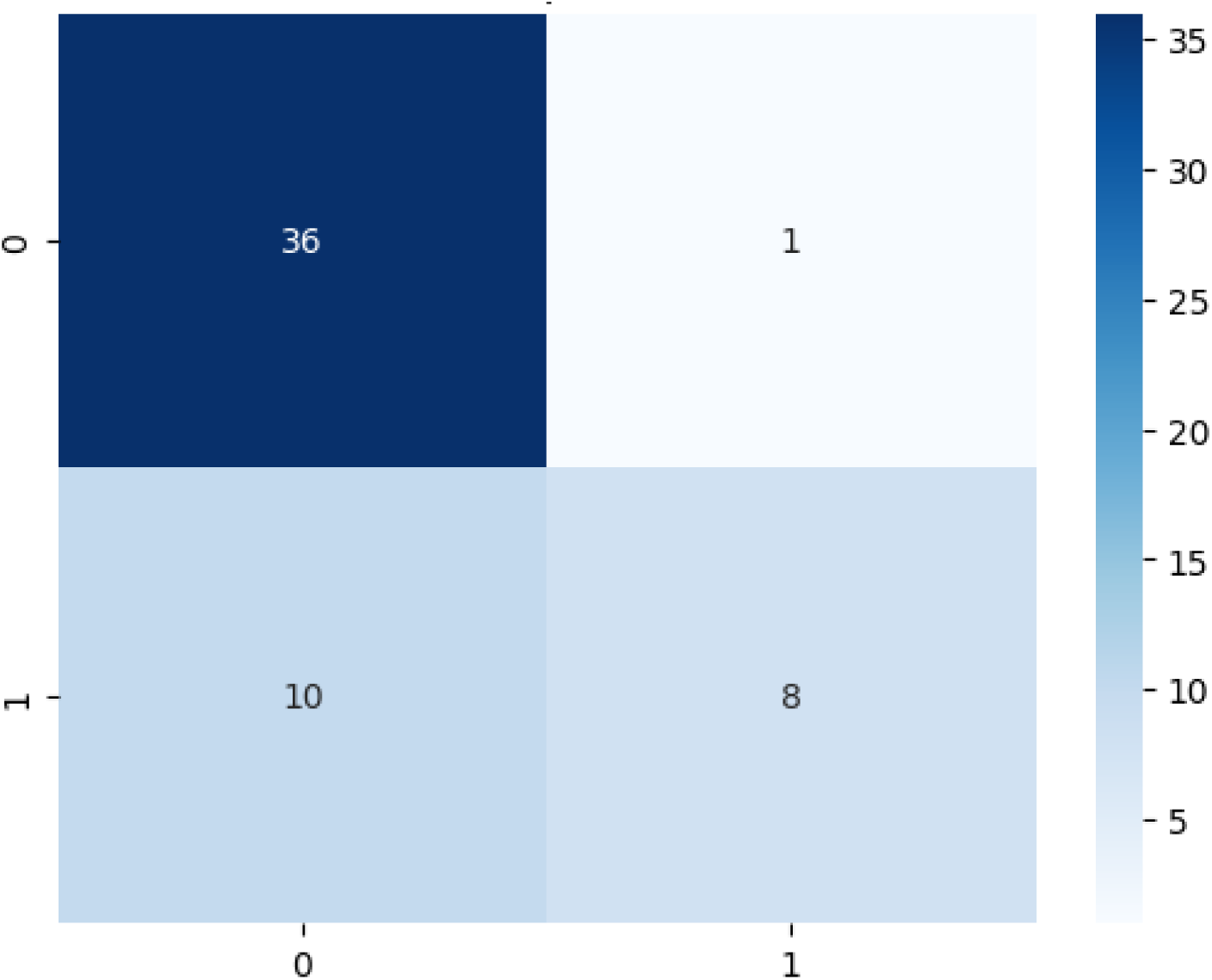
Confusion matrix of the text-based Random Forest model on the independent hold-out test set.

**Fig 2.**
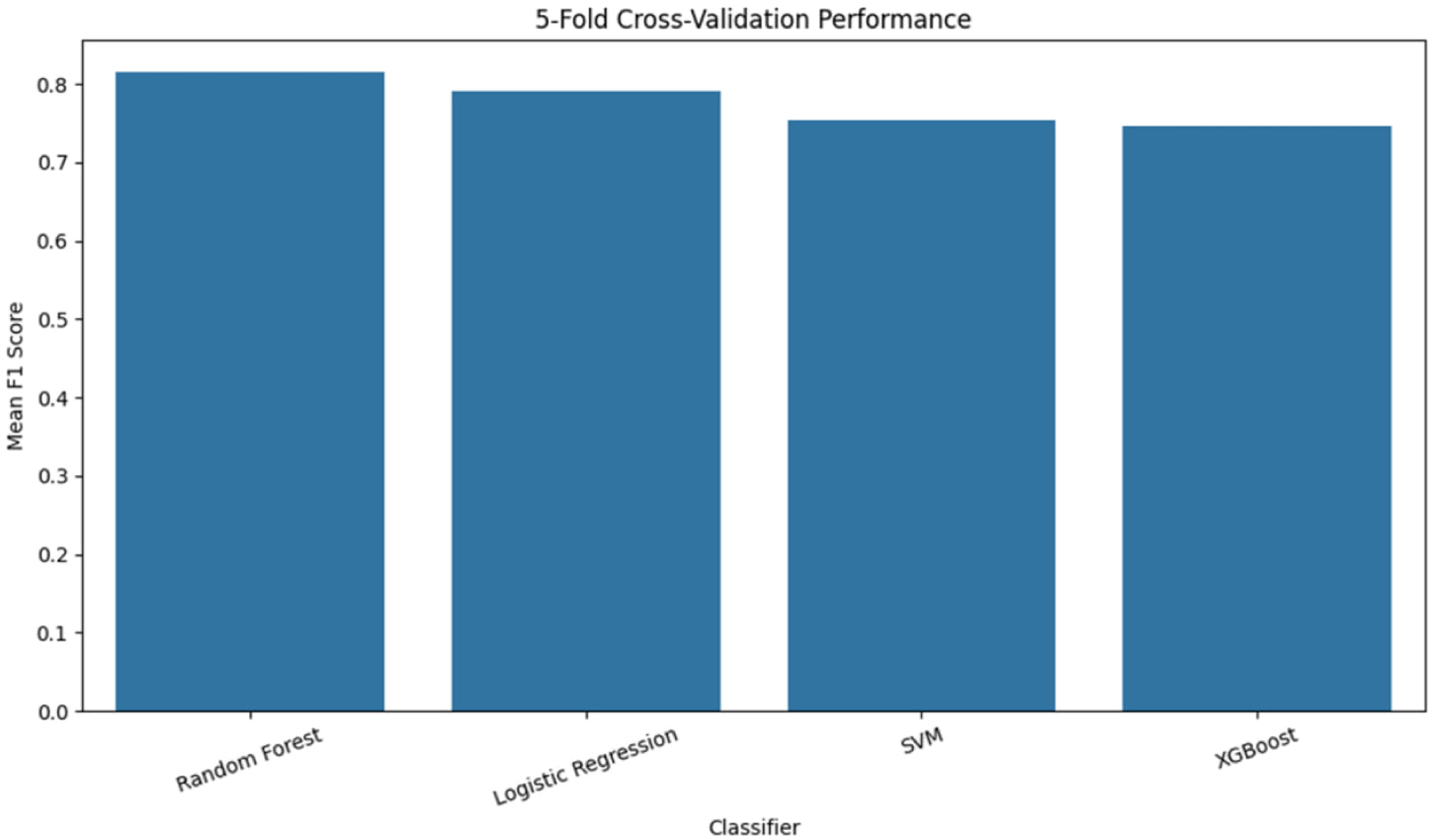
Comparison of text-based models using five-fold cross-validation. Error bars indicate standard deviation across folds.

**Fig 3.**
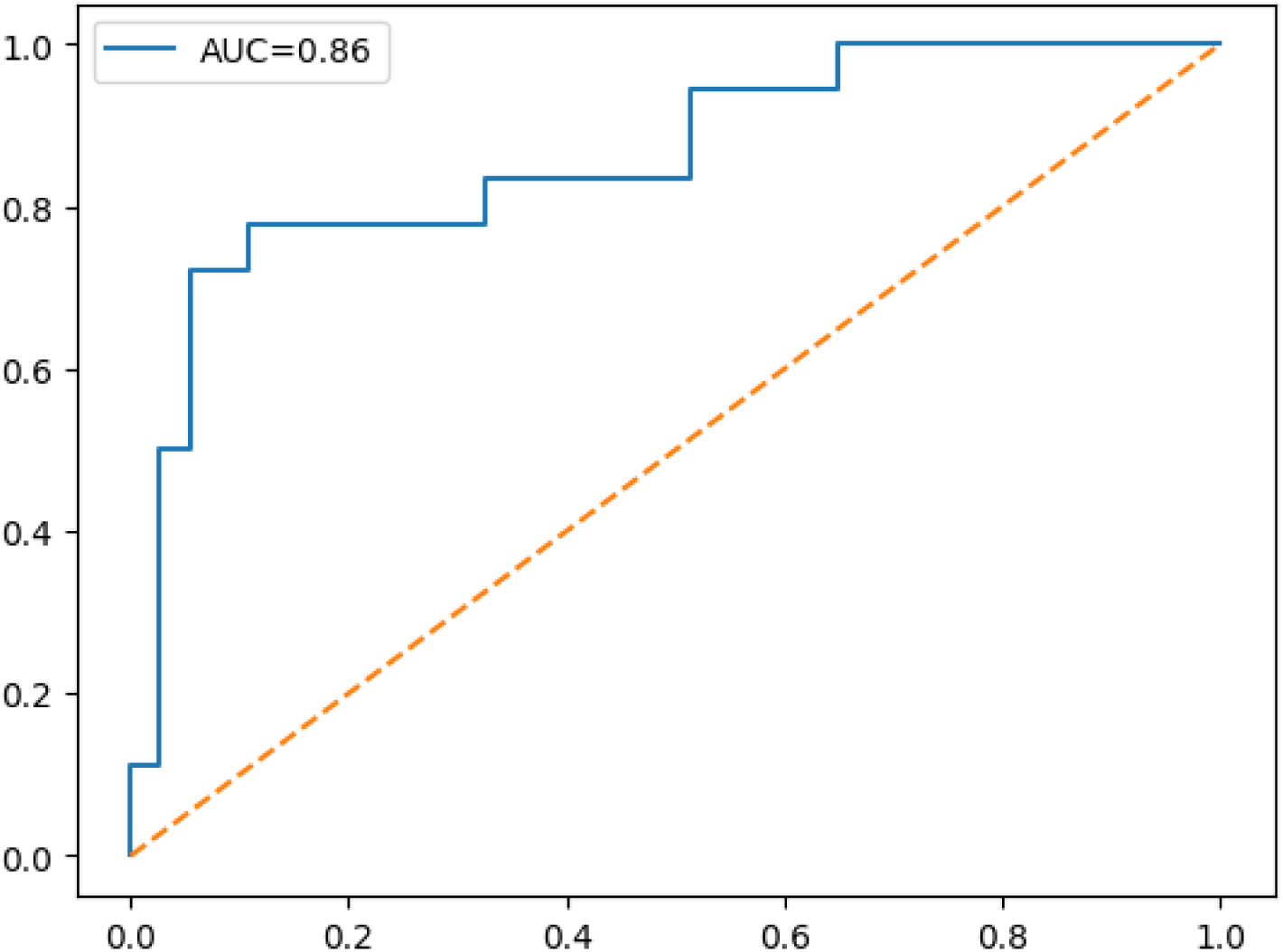
ROC curve of the text-based Random Forest model on the independent hold-out test set. The model achieved a ROC-AUC of 0.8673 (displayed rounded as 0.867).

**Fig 4.**
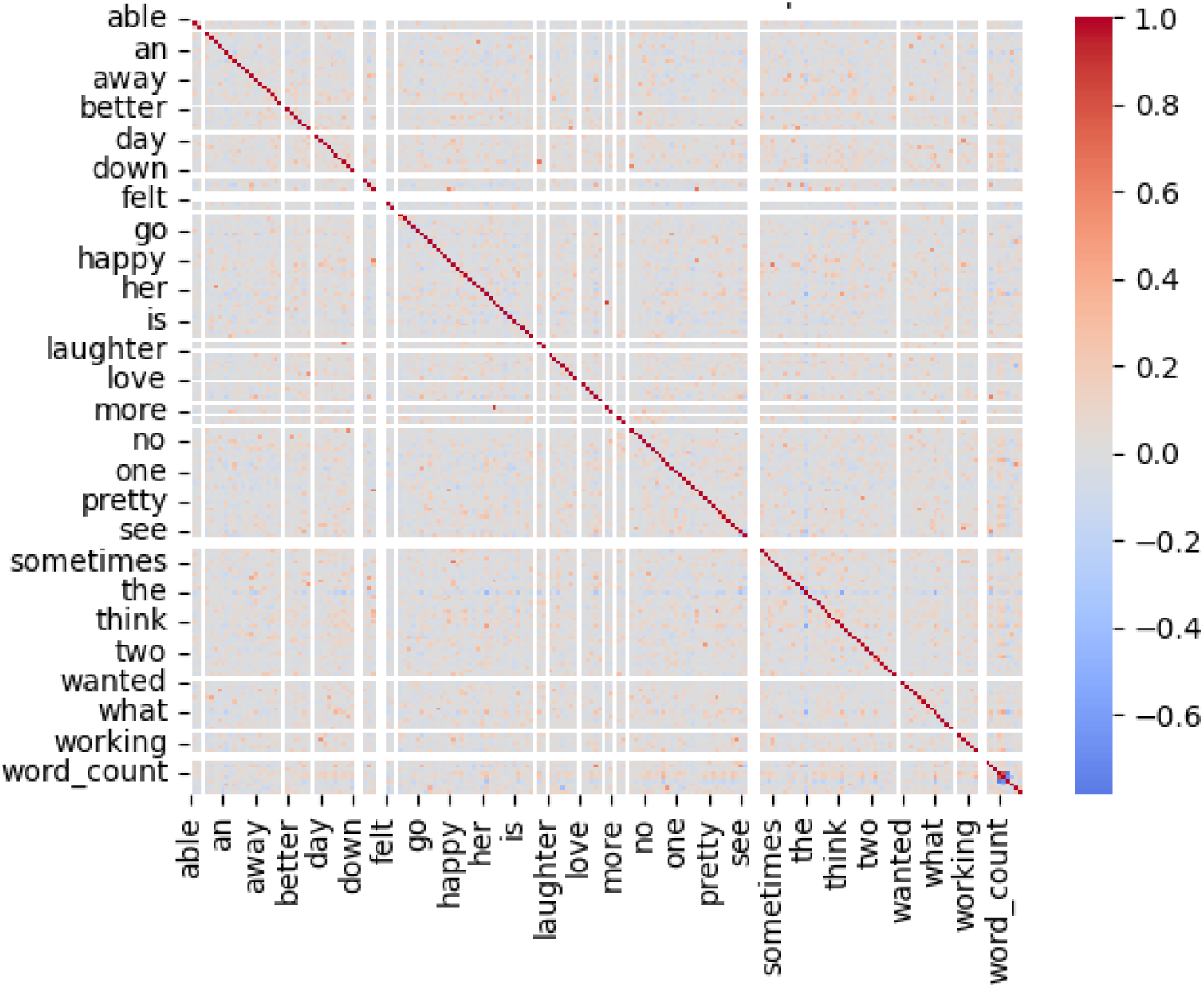
TF-IDF heatmap illustrating the distribution of term-weight values across selected textual instances. Darker cells represent higher TF-IDF weights.

**Fig 5.**
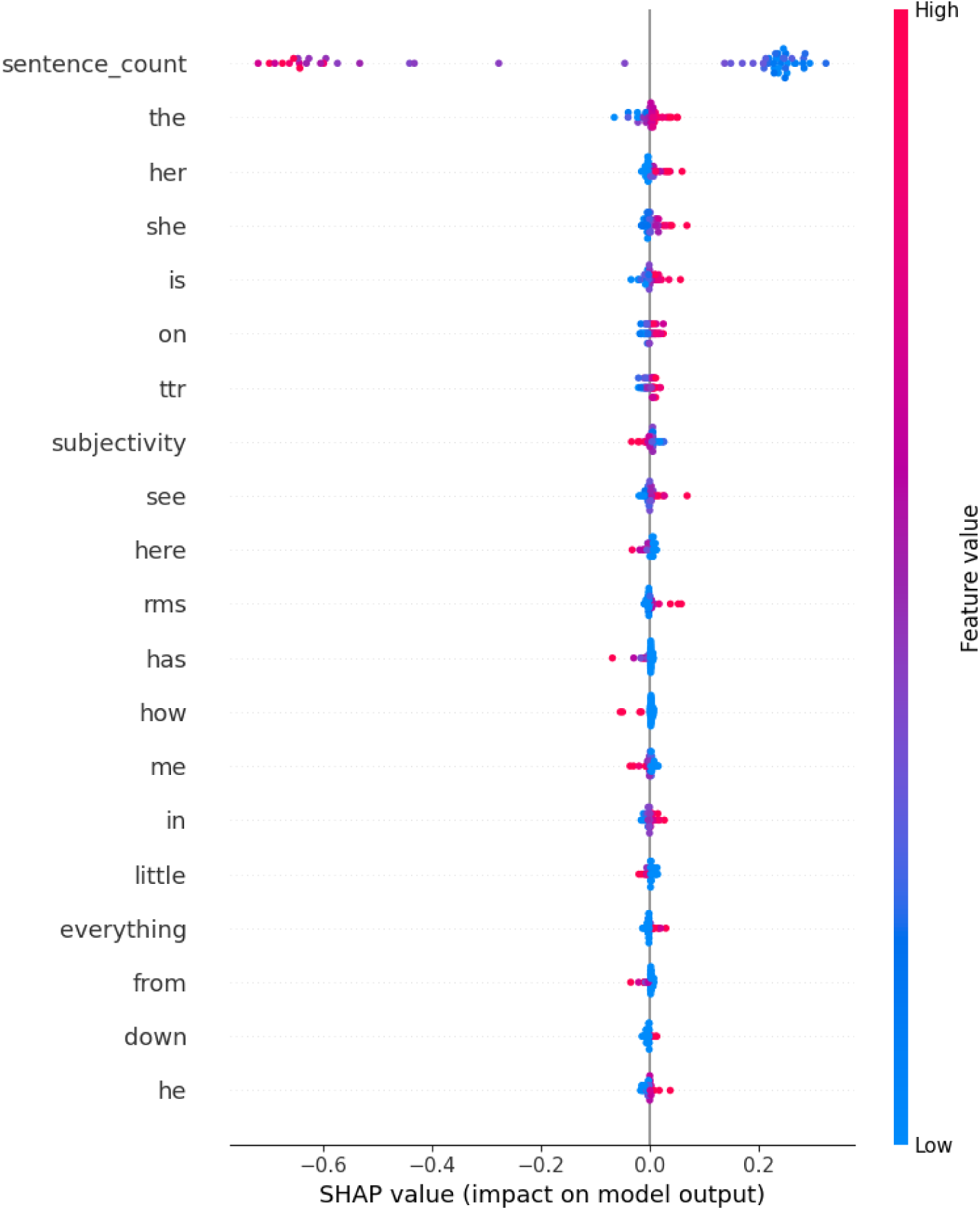
SHAP summary plot of the text-based model. Features are ranked according to their mean absolute SHAP contribution to the structural boundary.

**Table 3.**
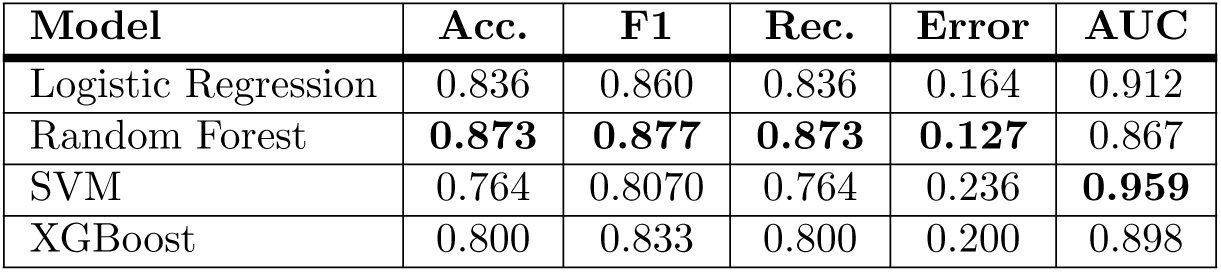
Performance of text-based structural validation models on the independent hold-out test set. Best values are shown in bold.

| Model | Acc. | F1 | Rec. | Error | AUC |
| --- | --- | --- | --- | --- | --- |
| Logistic Regression | 0.836 | 0.860 | 0.836 | 0.164 | 0.912 |
| Random Forest | **0.873** | **0.877** | **0.873** | **0.127** | 0.867 |
| SVM | 0.764 | 0.8070 | 0.764 | 0.236 | **0.959** |
| XGBoost | 0.800 | 0.833 | 0.800 | 0.200 | 0.898 |

Overall, the text-only results indicate that linguistic representations provide informative but relatively variable signals for isolating the target pseudo-labelled groups. Random Forest mapped the strongest overall classification boundary, whereas SVM showed the strongest ranking capability. These findings establish the textual baseline for the subsequent audio and multimodal structural evaluations.

### Audio-based experiment

The audio-only experiment used a 19-dimensional acoustic representation comprising RMS energy, zero-crossing rate (ZCR), spectral centroid, spectral roll-off, jitter, shimmer, and 13 Mel-frequency cepstral coefficients (MFCC_0_ through MFCC_12_). The training set contained 218 participants. To mitigate class imbalance and restrict the model from simply memorising the training distribution, the dataset was expanded to 356 instances using training-only augmentation. Specifically, because the acoustic latent space exhibited a more severe pseudo-label imbalance than the textual representation, Synthetic Minority Over-sampling Technique (SMOTE) was applied exclusively to the acoustic feature vectors of the minority class within each training fold. The number of synthetic observations generated was determined by the class distribution within that training fold; no synthetic observations were created from validation or test participants. The independent test set remained strictly unmodified at 55 original participants to ensure a rigorous evaluation. The target assignments were derived using the unsupervised procedure described in the methodology.

Key findings:

- **Best hold-out separability:** As detailed in Table 4, XGBoost achieved the highest test accuracy (90.91%) and F1-score (0.9061), with an error rate of 9.09%.
- **Best cross-validated model:** SVM achieved the highest mean accuracy (93.42 *±* 2.04%), F1-score (93.36 *±* 2.13%), recall (93.42 *±* 2.04%), and ROC-AUC (0.9761 *±* 0.0292), and was therefore selected as the final audio-based structural model. Its discriminative boundary on the independent test set is further illustrated by its confusion matrix (Fig 6) and ROC curve (Fig 7).
- **Model stability:** SVM showed remarkably low variability across folds, providing comparatively consistent performance across the cross-validation folds despite the higher single-point estimate obtained by XGBoost on the independent test set.
- **Interpretability:** The relationships among the extracted acoustic features are visualised in the correlation heatmap (Fig 8). Correspondingly, SHAP analysis (Fig 9) identified spectral roll-off and MFCC-related variables among the influential acoustic features, highlighting the heavy reliance on spectral and cepstral variance to geometrically divide the clusters.

**Fig 6.**
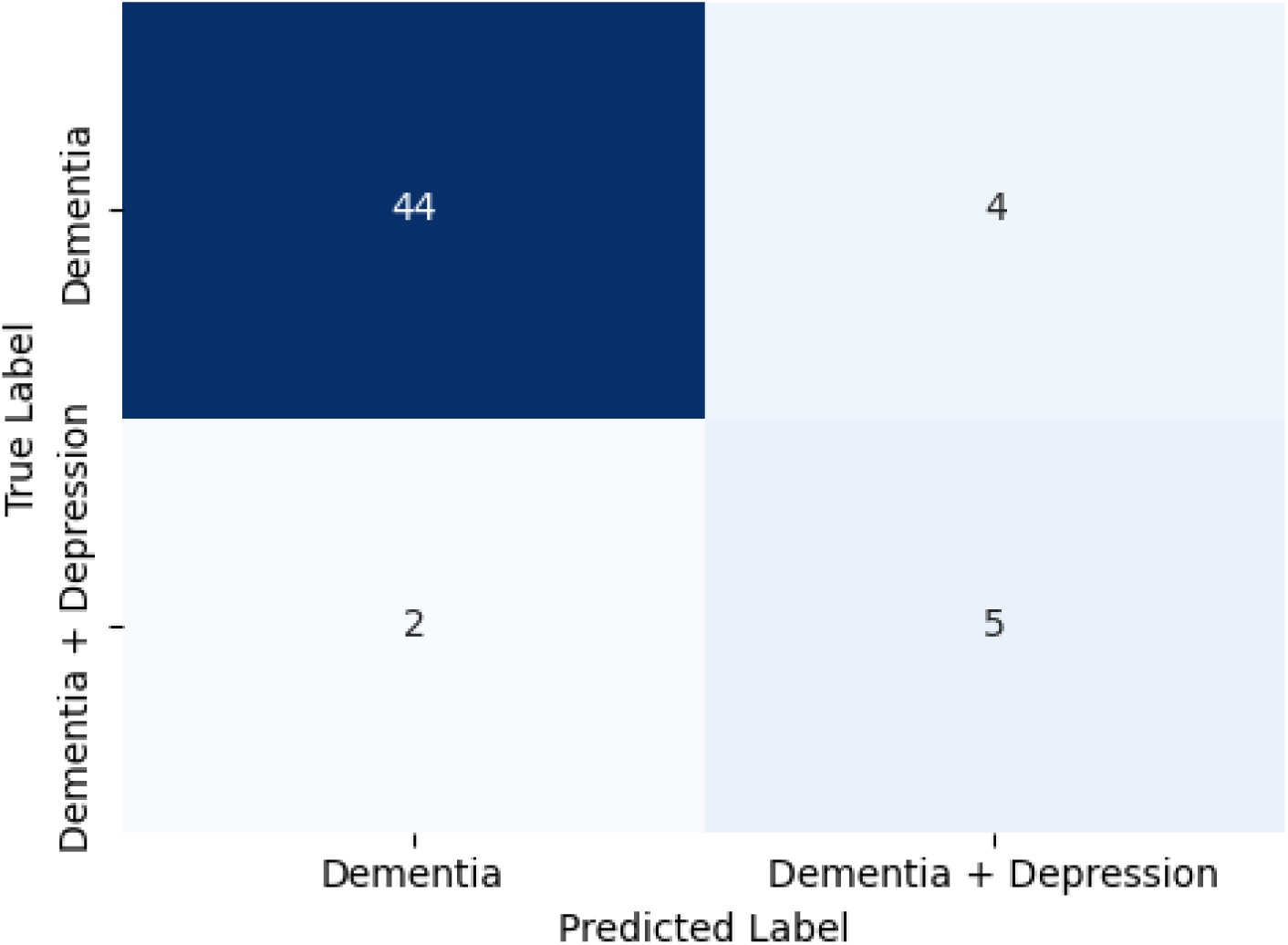
Confusion matrix of the audio-based SVM model for reproducing the representation-derived cluster assignments on the independent hold-out test set.

**Fig 7.**
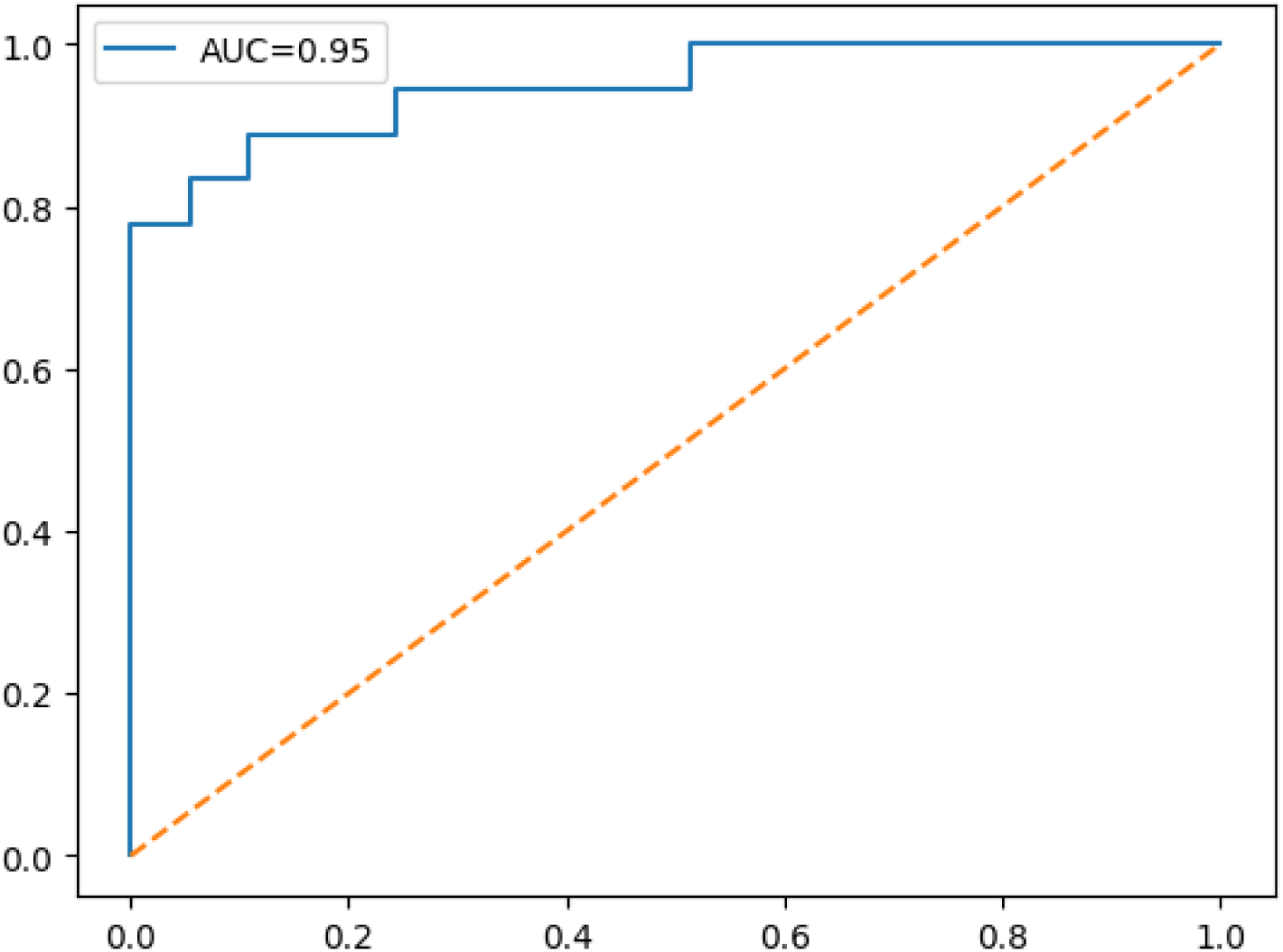
ROC curve of the audio-based SVM model on the independent hold-out test set. The model achieved a ROC-AUC of 0.9464 (displayed rounded as 0.95).

**Fig 8.**
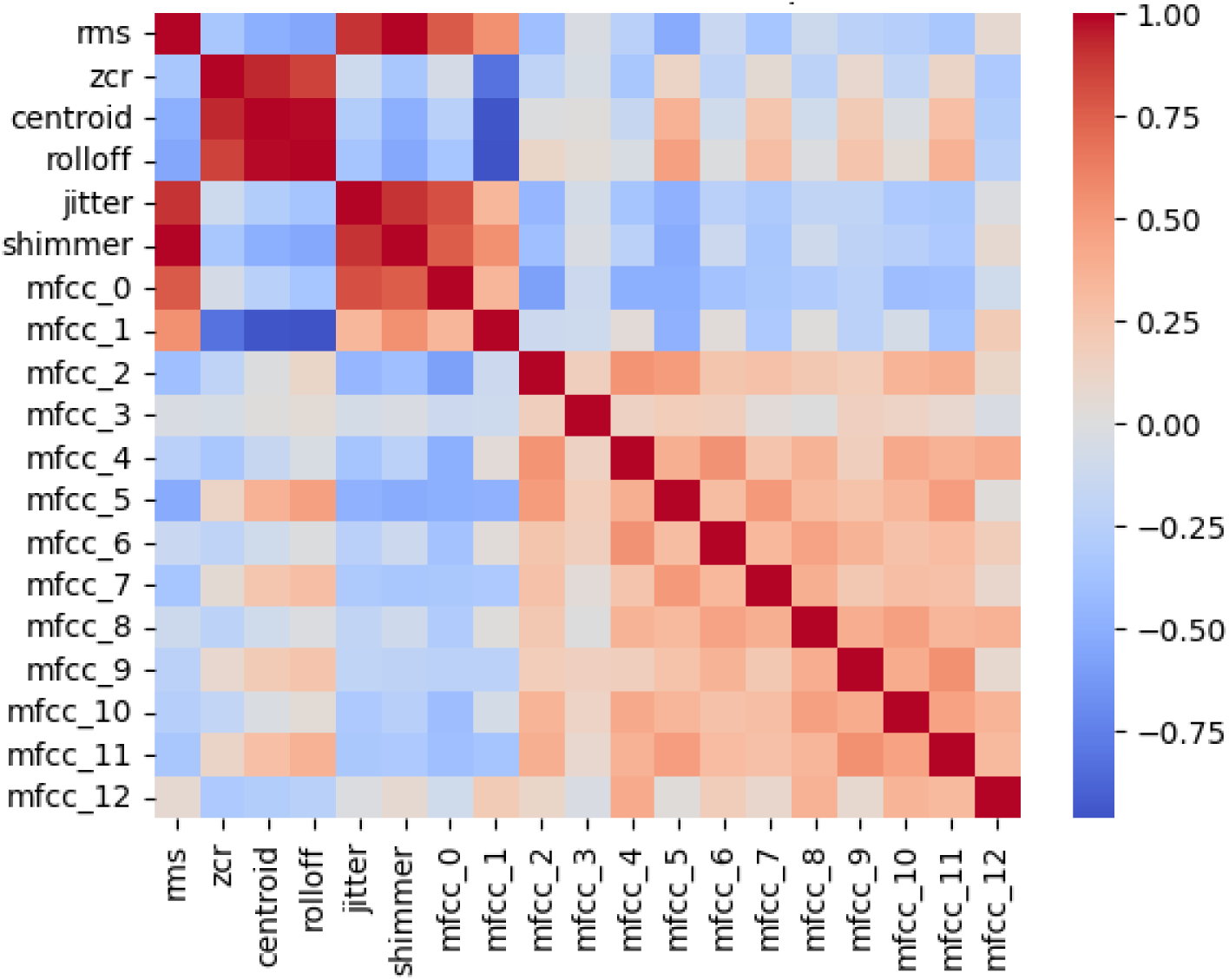
Correlation heatmap of the extracted acoustic features, illustrating relationships among RMS energy, zero-crossing rate, spectral descriptors, jitter, shimmer, and MFCC coefficients.

**Fig 9.**
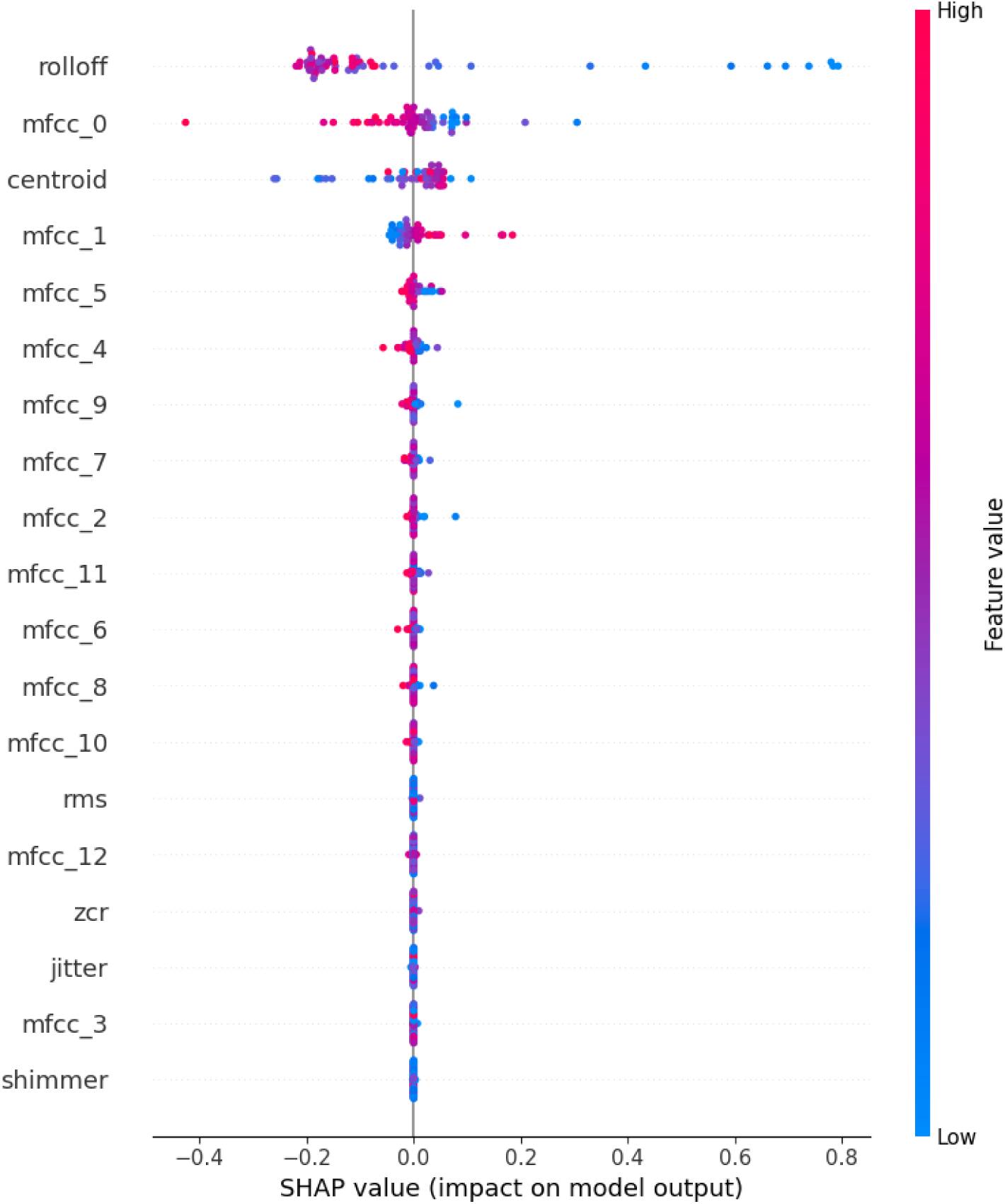
SHAP summary plot of the audio-based model. Features are ranked according to their mean absolute SHAP contribution, illustrating the relative geometric influence of acoustic variables on cluster separation.

**Table 4.** Performance of audio-based structural validation models on the independent hold-out test set. Best values are shown in bold.

| Model | Acc. | F1 | Rec. | Error | AUC |
| --- | --- | --- | --- | --- | --- |
| Logistic Regression | 0.818 | 0.839 | 0.818 | 0.182 | 0.8125 |
| Random Forest | 0.836 | 0.848 | 0.836 | 0.164 | 0.893 |
| SVM | 0.891 | 0.897 | 0.891 | 0.109 | 0.946 |
| XGBoost | **0.909** | **0.906** | **0.909** | **0.091** | 0.914 |

Overall, the audio representation provided a more consistently learnable and distinct basis for separating the discovered sub-phenotypes than the text-only representation. Although XGBoost achieved the highest single-test accuracy (Table 4), SVM provided the strongest cross-validated structural boundary and was therefore retained for multimodal comparison.

### Multimodal fusion experiment

The multimodal experiment combined the abstract acoustic and linguistic latent representations and evaluated four classical boundaries (Logistic Regression, Random Forest, SVM, and XGBoost) alongside the previously retained ANN-128 and ANN-256 neural configurations. The target groups were strictly the pseudo-labels generated via the unsupervised methodology; thus, performance represents algorithmic stability rather than clinically verified diagnosis.

Key findings:

- **High structural separability of the fused space:** As detailed in Table 5, the retained ANN-256 configuration achieved the highest multimodal cluster separability on the test set, with 92.73% accuracy, an F1-score of 0.9473, a macro-averaged recall of 0.9032, and an error rate of 7.27%, mathematically aligning with the structural threshold matrix. Although the SVM baseline produced a marginally higher AUC (0.9981), ANN-256 was explicitly selected as the retained model because accuracy and F1-score serve as our primary decision criteria for evaluating discrete structural separability. The high accuracy of this mapped structural boundary is further demonstrated by its confusion matrix (Fig 10) and ROC curve (Fig 11).
- **Classical-model baseline:** Among the linear and tree-based models, SVM provided the strongest five-fold cross-validation performance, achieving 89.41 *±* 5.38% accuracy and an F1-score of 89.37 *±* 5.38%.
- **Neural-model advantage:** As shown in Table 5, ANN-128 achieved 92.73% accuracy and an F1-score of 0.9307, while ANN-256 provided the strongest retained result. Within the two neural configurations evaluated, ANN-256 produced a higher F1-score than ANN-128 on the held-out test set. Given the small independent test set (*N* = 55), this minor variance in F1-score likely represents a threshold shift for a single participant rather than a definitive advantage of increased network capacity.
- **Complementary stability:** The multimodal representation produced the highest held-out separability among the evaluated representation-specific partitions. Because the target partitions were independently generated for each representation, this numerical difference should not be interpreted as a direct estimate of the incremental benefit of multimodal fusion.

**Fig 10.**
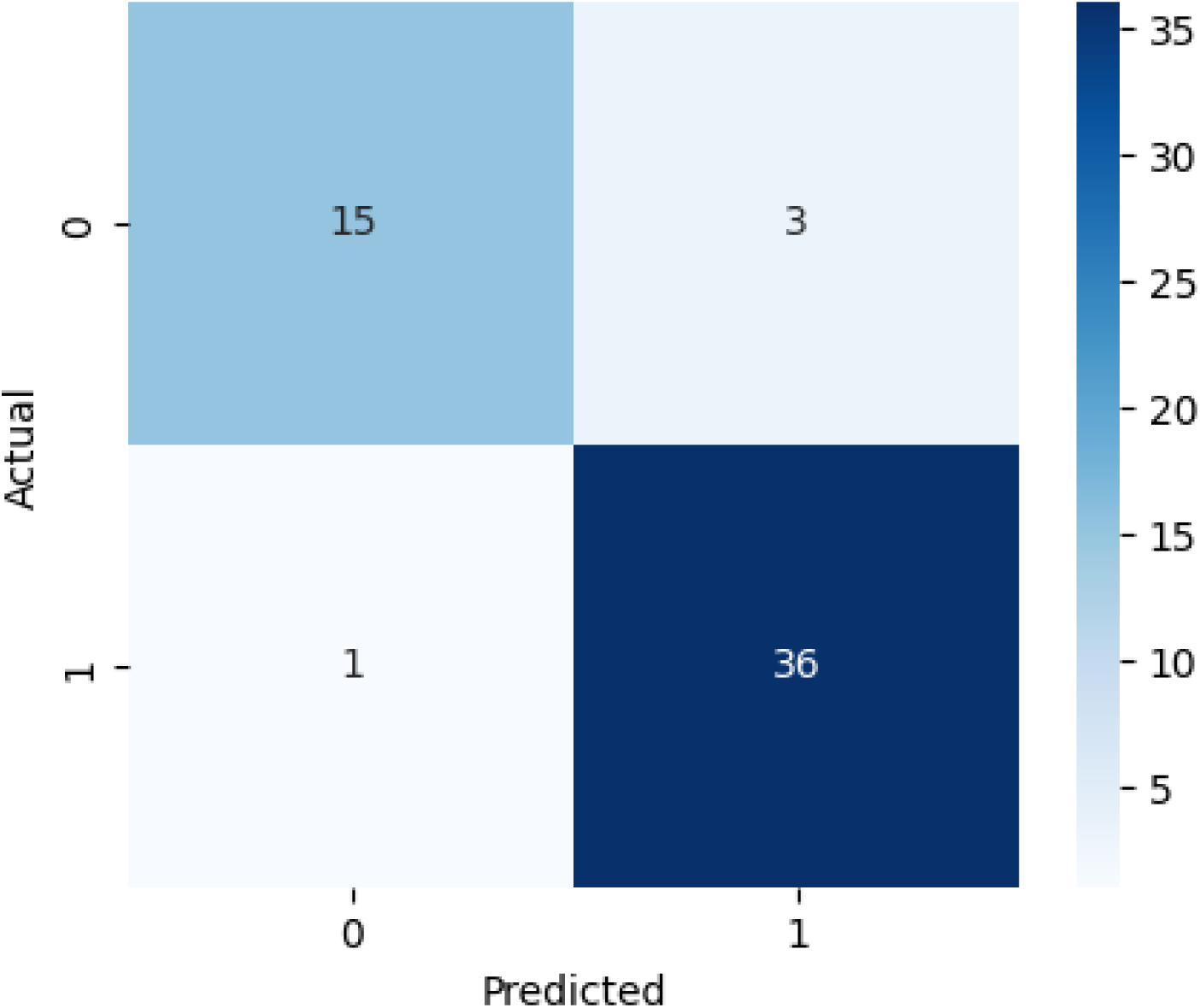
Confusion matrix of the retained multimodal ANN-256 model on the independent hold-out test set, demonstrating the high accuracy of the mapped structural boundary.

**Fig 11.**
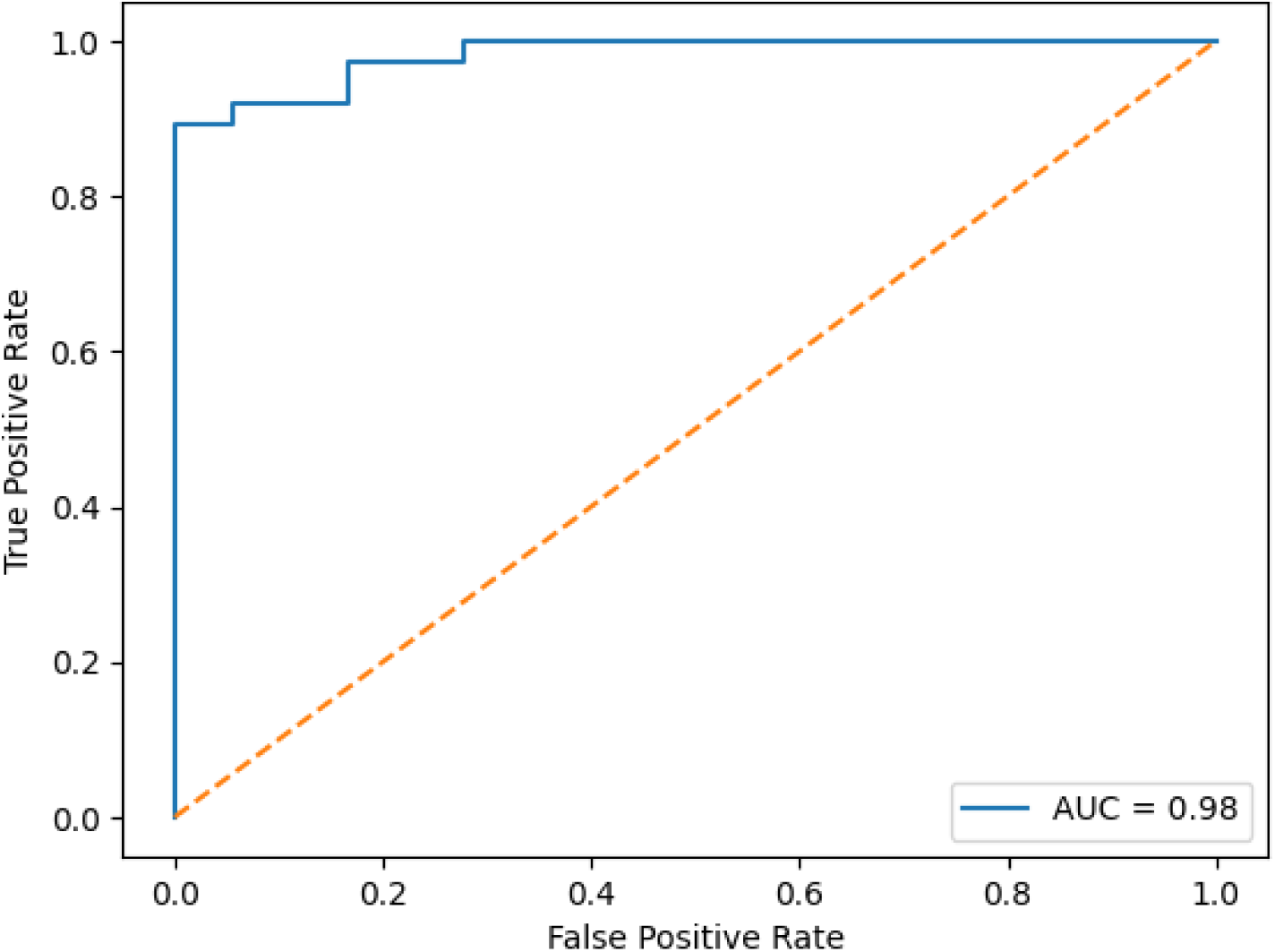
ROC curve of the retained multimodal fusion model on the independent hold-out test set. The model achieved a ROC-AUC of 0.9903 (displayed rounded as 0.99).

**Table 5.** Performance of multimodal structural validation models on the independent hold-out test set. Best values are shown in bold.

| Model | Acc. | F1 | Rec. | Error | AUC |
| --- | --- | --- | --- | --- | --- |
| Logistic Regression | 0.655 | 0.682 | 0.655 | 0.346 | 0.8953 |
| Random Forest | 0.873 | 0.882 | 0.873 | 0.127 | 0.996 |
| SVM | 0.891 | 0.898 | 0.891 | 0.109 | 0.998 |
| XGBoost | 0.873 | 0.882 | 0.873 | 0.127 | 0.990 |
| ANN-128 | 0.927 | 0.931 | 0.9273 | 0.073 | 0.988 |
| ANN-256 | **0.927** | **0.947** | **0.903** | **0.073** | **0.990** |

The multimodal experiment indicates that the two representation-derived target clusters were readily separable in the evaluated latent space. The cluster associated heuristically with the source depression representation should not be interpreted as a clinically confirmed depression subgroup. Achieving 92.73% test accuracy (Table 5) provides evidence that the representation-derived GMM partition was learnable from held-out participant data, actively avoiding the circularity of treating the supervised output as a clinical diagnostic score.

### Feature correlation and explainability analysis

To understand the structural drivers separating the discovered sub-phenotypes, the multimodal representation was analysed using feature visualisation and SHAP-based attribution. Rather than implying clinical causality, this analysis specifically identifies the mathematical boundaries learned by the model. The TF-IDF heatmap (Fig 12) illustrates the term-weight variance driving the textual component’s separability, highlighting specific lexical clusters that distinguish the subgroups. Correspondingly, the SHAP analysis (Fig 13) quantifies the relative magnitude of acoustic and linguistic features in defining the decision boundary of the retained multimodal model.

**Fig 12.**
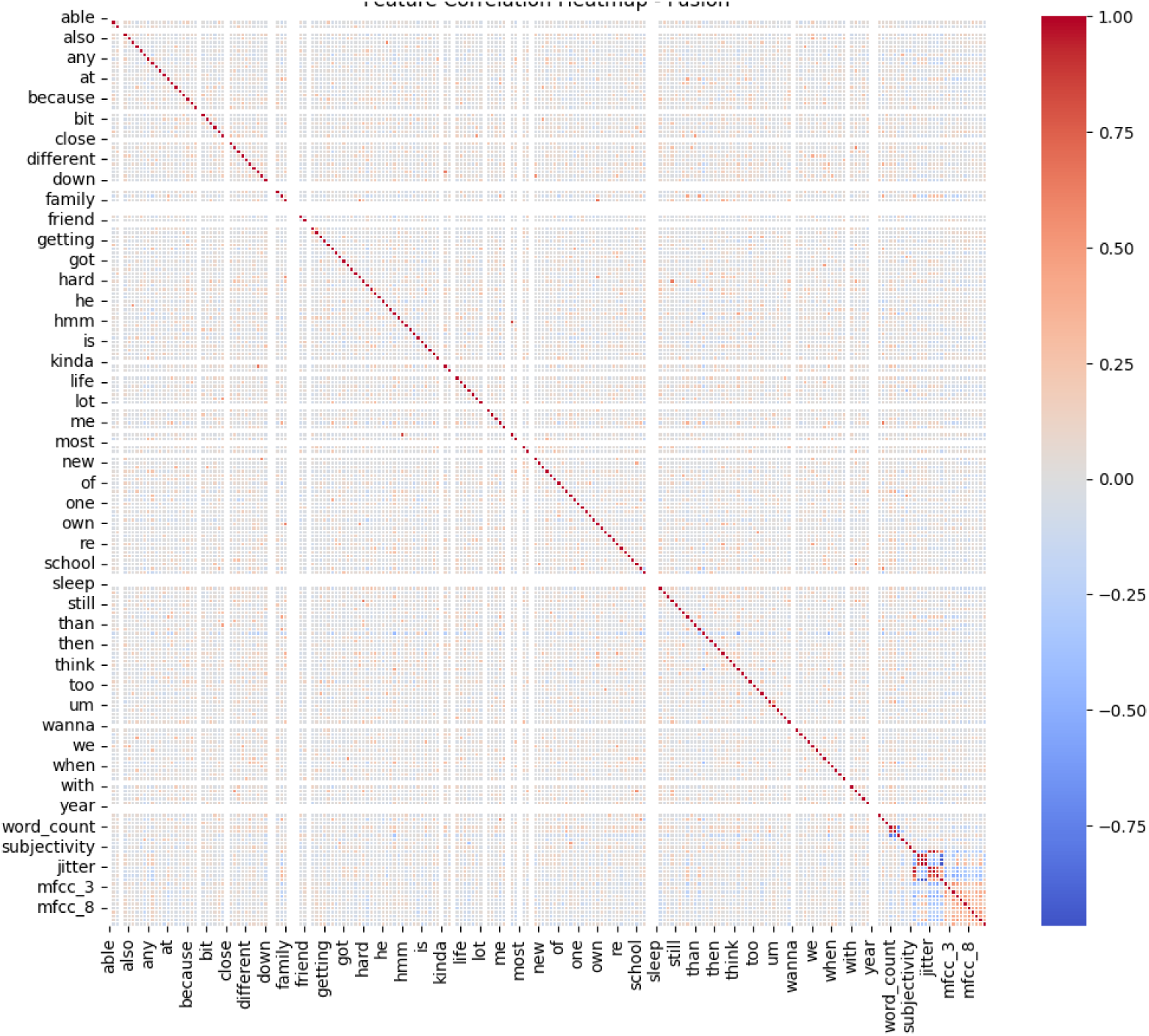
TF-IDF heatmap illustrating the distribution of term-weight values across selected textual representations used in the multimodal analysis. Darker cells correspond to higher TF-IDF weights.

**Fig 13.**
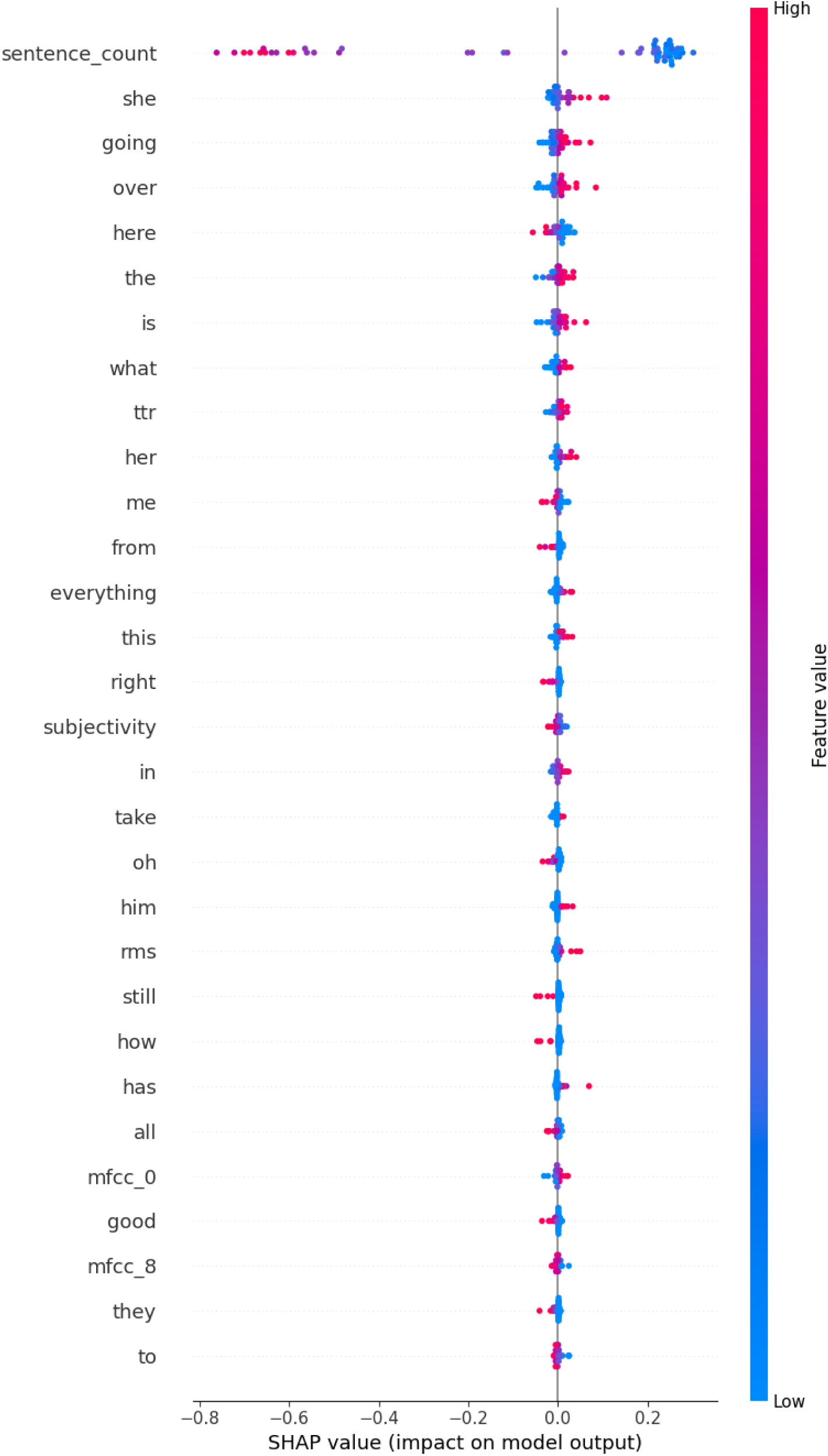
SHAP summary plot of the retained multimodal model. Features are ranked according to their mean absolute SHAP contribution, illustrating the relative mathematical influence of acoustic and linguistic variables on the decision boundary.

Key findings:

- **Modality-specific partitioning:** Audio achieved 89.09% accuracy (F1: 0.8966) on its specific pseudo-label target, while text achieved 87.27% (F1: 0.8770) on its respective target. Rather than indicating direct superiority, this highlights that the acoustic latent space formed a more easily learnable structural partition for its specific target distribution than the textual space did for its own.
- **Fusion characteristics:** The multimodal neural configurations mapped a distinct composite topology. The retained ANN-256 configuration achieved a high structural separation of 92.73% on the fused pseudo-labels. We note this evaluates the separability of the fused geometric space itself, rather than serving as an incremental gain over a static baseline, given the shifting class distributions (as evidenced by the differing confusion matrices).
- **SHAP interpretation:** Among the acoustic variables, spectral roll-off and several MFCC-related features had relatively large mean absolute SHAP contributions in the retained model, indicating that these variables contributed strongly to the model’s separation of the representation-derived groups, while sentence count, lexical diversity, and other linguistic features provided complementary geometric boundaries.
- **Interpretation boundary:** SHAP values here represent the model’s structural reliance on specific variables to separate the abstract sub-phenotypes; they strictly highlight geometric feature importance rather than independently validated candidate speech features.

The comparative overview in Table 6 summarizes the structural learnability across the three independent feature spaces. Because the pseudo-labels were generated independently for each space, these metrics represent how well models map each distinct modality’s topology, rather than an apples-to-apples incremental gain on a single, static diagnostic target. The SHAP analysis further suggests that the multimodal model maps its boundaries primarily on acoustic variance while incorporating selected linguistic characteristics.

**Table 6.**
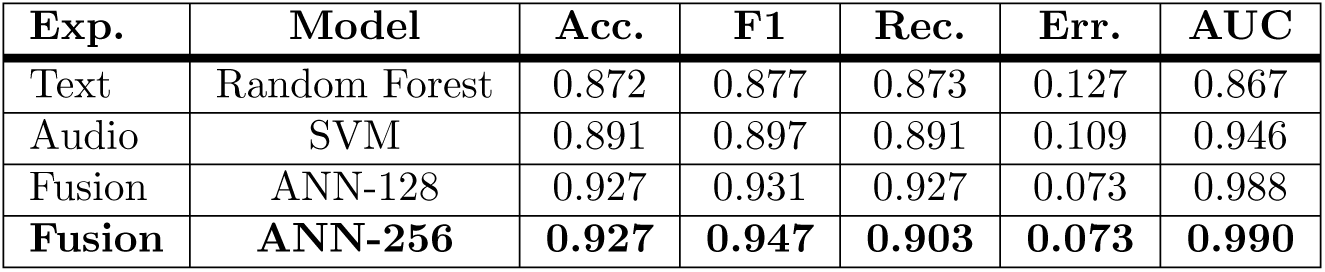
Comparison of text-only, audio-only, and retained multimodal experiments. Best values are highlighted in bold.

| Exp. | Model | Acc. | F1 | Rec. | Err. | AUC |
| --- | --- | --- | --- | --- | --- | --- |
| Text | Random Forest | 0.872 | 0.877 | 0.873 | 0.127 | 0.867 |
| Audio | SVM | 0.891 | 0.897 | 0.891 | 0.109 | 0.946 |
| Fusion | ANN-128 | 0.927 | 0.931 | 0.927 | 0.073 | 0.988 |
| **Fusion** | **ANN-256** | **0.927** | **0.947** | **0.903** | **0.073** | **0.990** |

As a final exploratory measure, the SHAP-derived Multimodal SHAP Attribution Magnitude (MSAM) was calculated using Eq 9. The MSAM summarises the raw magnitude of selected acoustic and linguistic SHAP contributions for an individual instance; it functions strictly as a model-attribution summary score, establishing a computational baseline for future prospective clinical validation.

## Discussion

This study provides an exploratory analysis of depression-associated speech representations within an Alzheimer’s disease (AD) cohort in the absence of independently observed depression labels. The principal finding was that the target-domain speech representations supported a two-cluster solution, with the *k* = 2 configuration yielding the highest Silhouette score among the candidate solutions evaluated. The subsequent supervised analyses showed that these representation-derived partitions were learnable from held-out participant data, with the retained multimodal ANN-256 model achieving 92.73% accuracy and an F1-score of 0.9473 on the independent test set. Acoustic representations also produced a highly learnable partition, with the retained SVM achieving 89.09% accuracy on its representation-specific test target. These results indicate that the discovered speech subgroups contain reproducible computational structure. They should not, however, be interpreted as evidence that one subgroup represents clinically diagnosed depression, because depression status was not independently established in the AD cohort. The present findings are broadly consistent with previous work showing that speech contains measurable linguistic and acoustic variation associated with depression and neurocognitive impairment, while also illustrating the difficulty of separating these phenomena in heterogeneous clinical populations. Fraser et al. [3] demonstrated that linguistic features could provide information relevant to depression in the context of Alzheimer’s disease, although the reported classification performance was substantially more modest than the structural separability observed in the present analysis. Similarly, Ehghaghi et al. [9] showed that data-driven clustering can reveal structure associated with depression and dementia from speech and language data. The present study extends this line of work by treating clustering primarily as a mechanism for discovering representation-derived subgroups rather than as a substitute for clinical diagnosis. Previous work using artificial neural networks to distinguish depression-related groups in AD and mild cognitive impairment also reported promising but limited performance [10]. The higher numerical values observed here therefore should not be interpreted as evidence of superior depression-detection performance, because the prediction target in the present study is an unsupervised cluster assignment rather than an independently assessed clinical outcome. The relatively strong contribution of acoustic variables provides a plausible methodological explanation for the observed structure, although the present design does not establish a specific physiological or affective mechanism. Spectral roll-off, MFCC-related variables, and other acoustic descriptors may capture differences in vocal energy distribution, spectral characteristics, articulation, and voice quality that are not fully represented by lexical features. This is particularly relevant in AD, where changes in speech production and connected language can arise from cognitive, motor, and communicative factors. The SHAP analysis showed that several acoustic variables contributed strongly to the learned decision boundary, while sentence count, lexical diversity, and other linguistic variables contributed additional information. Importantly, these attributions describe the variables used by the computational model to reproduce the representation-derived partition; they should not be regarded as independently validated biomarkers of depression. The stronger acoustic separability may therefore reflect a combination of affective variation, cognitive impairment, task-specific behaviour, recording characteristics, and other domain-related factors. Several limitations define the interpretation of these findings. First, the target AD cohort lacked independently assessed depression status; consequently, the cross-corpus alignment cannot establish depression prevalence, diagnostic accuracy, or clinical severity. Second, DementiaBank and DAIC-WOZ differ substantially in speech elicitation, participant characteristics, recording environment, and conversational structure. Although the feature spaces were constrained to permit geometric comparison, these domain differences may contribute to the observed alignment and cannot be separated from affective variation using the present design. Third, the independent test set comprised 55 participants, limiting the precision of performance estimates and making small differences between models difficult to interpret. Fourth, the pseudo-labels were generated independently within the text, acoustic, and fused representations, which inherently prevents a direct one-to-one comparison of modality-specific accuracy. Furthermore, to mitigate severe class imbalance within the acoustic latent space, SMOTE was applied exclusively to the audio training partitions. Consequently, the strong structural separability observed in the audio-only experiment may partially reflect the algorithmic benefits of this minority over-sampling, rather than an intrinsic superiority of acoustic features over textual representations. Finally, the SHAP-derived MSAM is currently a study-specific attribution summary. In future prospective trials, this single scalar value could potentially be utilised as a thresholding variable to trigger formal clinical referrals or additional psychiatric screening for patients exhibiting high depression-associated speech variance. Future studies should therefore evaluate the framework in prospectively recruited AD cohorts with independently assessed depression status, pre-specified clinical thresholds, larger and demographically diverse samples, and external validation cohorts. Such studies should test whether the representation-derived features remain associated with depression after accounting for cognitive severity, age, sex, medication, speech task, recording conditions, and other clinically relevant confounders.

## Conclusion

This study establishes an exploratory framework for examining depression-associated speech representations in Alzheimer’s disease when independently assessed depression labels are unavailable. The findings demonstrate that representation-derived speech subgroups can be reproduced computationally from held-out participant data, while the cross-corpus alignment provides a hypothesis-generating link to depression-associated speech variation.

The acoustic representation yielded higher held-out separability for its representation-specific cluster assignment than the corresponding text representation. Because the target partitions were independently generated, this difference should be interpreted as representation-specific learnability rather than evidence that acoustic features are intrinsically superior for detecting depression. The fused representation achieved the highest held-out separability among the evaluated representation-specific partitions. This finding is consistent with the possibility that acoustic and linguistic information provide complementary signals, although the independent pseudo-label generation prevents a direct estimate of their incremental clinical value. SHAP-based analysis successfully mapped the decision boundaries of these sub-phenotypes, identifying influential multimodal features. Furthermore, the proposed biomarker taxonomy and MSAM were established as exploratory, model-derived summaries of these feature attributions.

Overall, these findings provide preliminary evidence that multimodal speech representations can be used to explore representation-derived subgroups within an Alzheimer’s disease cohort and to generate hypotheses about behavioural speech variation. By explicitly measuring algorithmic stability rather than diagnostic efficacy, the framework provides an exploratory computational baseline for future clinically labelled studies. Future work must prioritise independent clinical labels, external cohorts, and rigorous prospective testing to translate these structural findings into validated clinical diagnostic tools.

## Data availability statement

The DAIC-WOZ dataset is available to researchers upon signing a Data Use Agreement via the University of Southern California. DementiaBank is accessible to affiliated researchers via TalkBank. Due to ethical restrictions regarding sensitive human speech recordings, the raw audio cannot be deposited in an unrestricted public repository.

## Code availability

All code developed by the authors for preprocessing, feature extraction, representation learning, clustering, classification, evaluation, and explainability will be made available in a public repository upon publication.

The repository will include the software environment, configuration files, random seeds, and instructions required to reproduce the reported analyses.

---

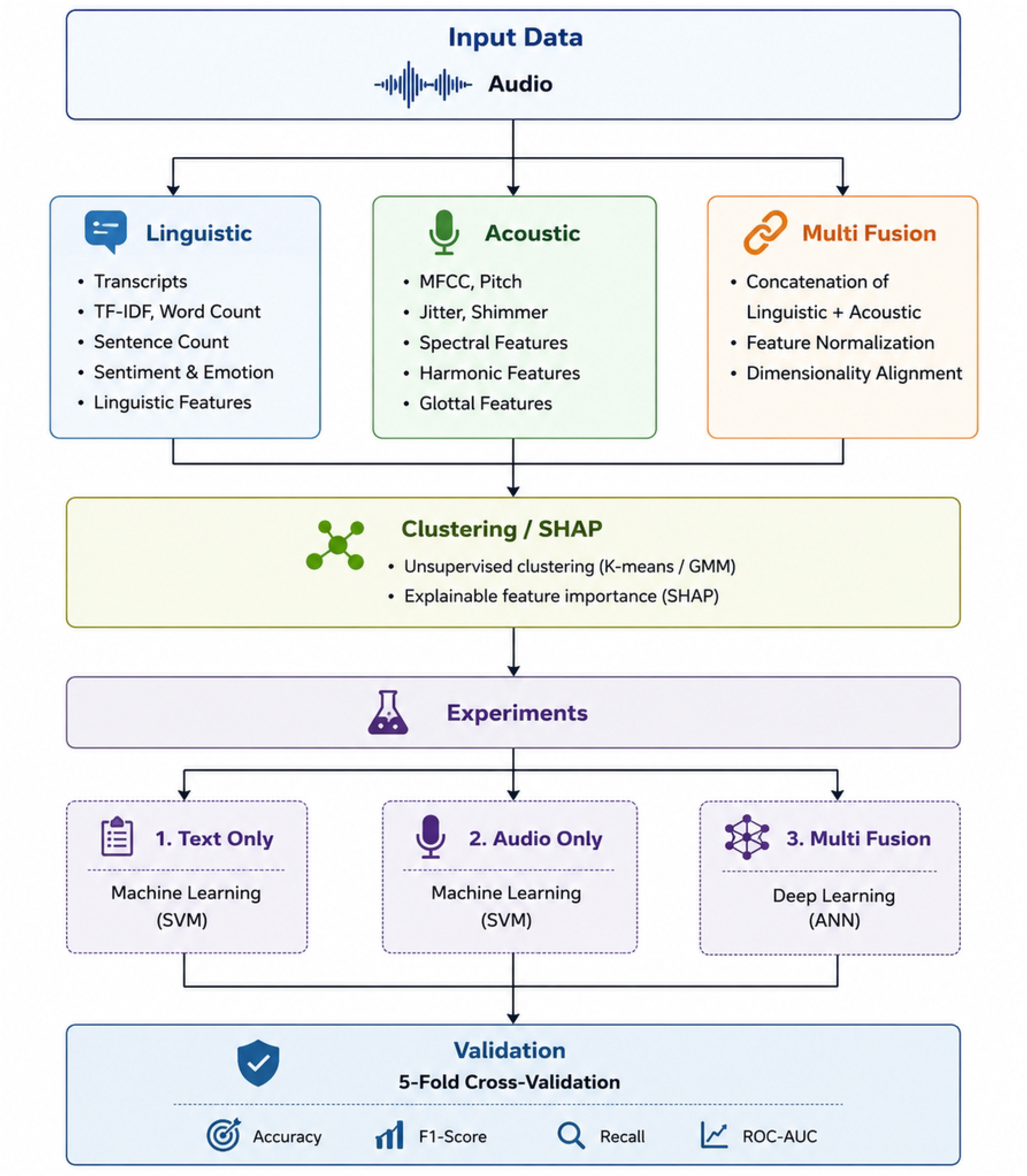
Main figure of the Multimodel.

